# Lower activity of cholesteryl ester transfer protein (CETP) and the risk of dementia: a Mendelian randomization analysis

**DOI:** 10.1101/2023.11.03.23298058

**Authors:** Amand F Schmidt, Michael H Davidson, Marc Ditmarsch, John J. Kastelein, Chris Finan

**Author notes:** Corresponding author: A Floriaan Schmidt.

## Abstract

Elevated levels of low-density lipoprotein cholesterol (LDL-C) are linked to dementia risk, and conversely, increased plasma concentrations of high-density lipoprotein cholesterol (HDL-C) and apolipoprotein-A1 (Apo-A1) associate with decreased dementia risk. Inhibition of cholesteryl ester transfer protein (CETP) meaningfully affects the concentrations of these blood lipids and may therefore provide an opportunity to treat dementia. Drug target Mendelian randomization (MR) was employed to anticipate the on-target effects of lower CETP concentration (µg/mL) on plasma lipids, cardiovascular disease outcomes, Lewy body dementia (LBD) as well as Parkinson’s dementia. MR analysis of lower CETP concentration recapitulated the blood lipid effects observed in clinical trials of CETP-inhibitors, as well as protective effects on CHD (odds ratio (OR) 0.92, 95% confidence interval (CI) 0.89; 0.96), heart failure, abdominal aortic aneurysm any stroke, ischemic stroke, and small vessel stroke (0.90, 95%CI 0.85; 0.96). Consideration of dementia related traits indicated that lower CETP concentrations were associated higher total brain volume (0.04 per standard deviation, 95%CI 0.02; 0.06), lower risk of LBD (OR 0.81, 95%CI 0.74; 0.89) and Parkinson’s dementia risk (OR 0.26, 95%CI 0.14; 0.48). *APOE4* stratified analyses suggested the LBD effect was most pronounced in *APOE-*ε4+ participants (OR 0.61 95%CI 0.51; 0.73), compared to *APOE-*ε4-(OR 0.89 95%CI 0.79; 1.01); interaction p-value 5.81×10^-4^. Additionally, MR was employed to link plasma CETP concentration to the levels of cerebrospinal fluid and brain proteins previously implicated in neurodegenerative pathways These results suggest that inhibition of CETP may be a viable strategy to treat dementia.

## Introduction

Cholesteryl ester transfer protein (CETP) facilitates the exchange of triglycerides (TG) and cholesterol ester between high-density lipoprotein cholesterol (HDL-C) and apolipoprotein-B (Apo-B) rich particles such as low-density lipoprotein cholesterol (LDL-C). CETP-inhibition has shown to elicit a plethora of beneficial effects on lipid metabolism, robustly decreasing the plasma concentration of canonical atherosclerosis particles such as total and small LDL, and lipoprotein (a) (Lp[a]), while increasing plasma concentrations of mature HDL, as well as pre-beta HDL and apolipoprotein-A1 (Apo-A1)^1–4^.

The four CETP-inhibitors (CETPi) evaluated in phase 3 clinical trials (anacetrapib, evacetrapib, dalcetrapib, torcetrapib) showed heterogenous effects on the magnitude of lipid perturbation, with an HDL-C percentage increase between 29% for dalcetrapib and ∼130% for anacetrapib/evacetrapib, and an LDL-C decrease between 1% for dalcetrapib and 20% for anacetrapib/evacetrapib^5^. This resulted in an equally mixed clinical effects profile^5^, with only the REVEAL trial for anacetrapib showing a non-HDL-C proportional protective effect of CETPi on CVD onset (rate ratio 0.91; 95% confidence interval (95%CI) 0.85; 0.97)^6^. The presence of meaningful differences in clinical effects profile strongly suggests that previous CETPi failures are likely attributable to the specific compound rather than to CETP inhibitors as a class^7^.

We have previously determined the viability of a reduction in CETP concentration using Mendelian randomization (MR), leveraging genetic instruments strongly associating with plasma CETP concentration, finding that lower plasma CETP concentration decreased the risk of CHD, heart failure (HF) and chronic kidney disease^5,8^. Because genetic variants are protected against confounding bias and reverse causation, MR provides a robust indication of the likely on-target effects of sufficiently potent drug target perturbation using data from human subjects^9–11^. A further benefit of MR is that it can utilize aggregated genetic data (e.g., variant-specific point estimates and standard errors) from independent studies to maximize the available sample size and hence precision.

Given the robust LDL-C lowering effects of CETP-inhibition, research has understandably focussed on its potential implications for cardiovascular disease (CVD) prevention. Multiple lines of evidence however suggest lipid metabolism is also involved with bioenergetic decline and chronic neuro-inflammation in the brain, which may contribute to neurodegenerative disorders^12^. This interrelationship between metabolism and neurodegeneration is further illustrated by the connection between amyloid-β, apolipoprotein-E (Apo-E) isoforms and lipid trafficking associating with the onset dementias such as Alzheimer’s disease (AD), Lewy body dementia (LBD), and dementia associated with Parkinson’s Disease (PD)^13–16^. Noting that LBD and dementia in PD are closely related diseases, both caused by underlying Lewy body disorders, which predominantly differ in temporal sequence of symptoms and clinical features^17^.

The Apo-E isoform *APOE-*ε4 is a major determinant of dementia risk, with homozygote carriers (*APOE-*ε4ε4) having an up to 15 fold increased risk^18,19^. Dementia in *APOE-*ε4 carriers^12^ is characterized by insufficient lipidation of Apo-E HDL particles, which dysregulates the fine balance between cholesterol availability to neurons and cholesterol accumulation in astrocytes, which has cytotoxic and proinflammatory consequences^20^. The lack of lipidation of Apo-E/HDL additionally affects astrocyte membrane composition, stimulating the formation of β-amyloid containing plaques, which is a major characteristic of AD brains. *APOE-*ε4 carriership and cholesterol metabolism has additionally been implicated in the development amyotrophic lateral sclerosis (ALS) as well as multiple sclerosis (MS)^21–23^.

Given the central role of CETP in HDL metabolism and the fact CETP inhibition raises plasma concentrations of both Apo-A1 and Apo-E^24^, we sought to elucidate a potential causal relationship between lower plasma CETP concentration and the risk of dementia and neurodegenerative diseases using MR. Specifically, we considered GWAS on LBD combined and stratified by *APOE-*ε4 status, as well as PD, dementia in PD – representing a disease clustering with a strong *APOE-*ε4 contribution. Furthermore, we considered potential associations with MS and ALS. As a positive control we first sought to confirm our previously reported effects on CHD, expanding this to additional CVD outcomes including ischemic stroke and small vessel stroke, and abdominal aortic aneurysm (AAA). We used *cis*-MR to identify associations between plasma CETP concentration and protein values in cerebrospinal fluid (CSF) and brain tissue, identifying associations with proteins previously implicated with neurodegeneration. Finally, we sought to replicate associations with biomarkers and disease onset by performing additional MR analyses weighting the *cis*-acting CETP variants by their association with Apo-A1, and Apo-B (downstream proxies of CETP activity)^25^. Importantly, as shown by Schmidt *et al.*^9,11^ a *cis*-MR analysis weighted by downstream effects of the protein does not require, or imply, that the weighting factor itself causes disease. Instead, the weights merely function as a proxy for protein value and activity. Hence inference in these Apo-A1 and Apo-B weighted analyses remains on the effect of lower CETP and does not address questions on potential lipoprotein mediation.

## Methods

### Selection of genetic instruments to model CETP effects

Genetic instruments associating with CETP concentration (µg/mL) were identified from a GWAS conducted by Blauw *et al.*^26^ To limit the potential for bias-inducing pre-translational horizontal pleiotropy^9,11^ we applied a *cis* window of ±25 kilobase pair (kbp) around *CETP* (ENSG00000087237, build 37), noting that this includes the entire GWAS signal observed by Blauw *et al*. Variants were selected to have an F-statistic of 24 or larger, and a minor allele frequency (MAF) of 0.01 or larger. The F-statistic was selected to limit the potential influence of weak-instrument bias^27^. The MAF threshold was chosen to ensure we could robustly model genetic linkage disequilibrium (LD)^9^ based on a random sample of 5,000 UK biobank participants as a reference. Using these references data, the genetic variants were clumped to an R-squared of 0.30, using the same reference data to model the residual LD (see below).

As described in Schmidt *et al.* 2020^9^ genetic associations with downstream consequences of protein expression can be used as an additional source of instruments selection and modelling using Mendelian randomization (MR). This provides opportunities to replicate the results observed in *cis*-MR using genetic associations with protein concentration. MR analyses using downstream proxies of protein expression reflect effects of protein activity, complementing analyses of protein level. Here we used GWAS^25^ on plasma concentration of Apo-A1 and Apo-B from which we extracted genetic variants, applying the same variant selection criteria centred on the *cis*-CETP region. We differentiate between the three analysis by referring to MR analyses weighted by “CETP”, “Apo-A1”, or “Apo-B”.

### Mendelian randomization analysis

*Cis-*MR was employed to ascertain the possible causal effects decreased CETP concentration on neurodegenerative disease and cardiovascular outcomes. MR estimates were calculated using generalized least squares (GLS) implementations of the inverse-variance weighted (IVW) estimator and the MR-Egger estimator, the latter being unbiased in the presence of horizontal pleiotropy at the cost of lower precision^28^. We used GLS to directly model the LD reference structure, after clumping to an R-squared of 0.30, optimizing power while preventing potential multicollinearity-based numerical instability^29^. To minimize the potential influence of horizontal pleiotropy, variants beyond 3 times the mean leverage or with an outlier (Chi-square) statistic larger than 10.83, were pruned^30^. Finally, a model selection framework was applied to select the most appropriate estimator, IVW or MR-Egger^30,31^. This model selection framework^32^ utilizes the difference in heterogeneity between the IVW Q-statistic and the Egger Q-statistic to decide which method provides the best model to describe the available data and hence optimizes the bias-variance trade-off.

### Involvement of plasma CETP with CSF and brain proteins implicated with neurodegeneration

We additionally conducted *cis*-MR analysis to identify associations between plasma CETP concentration and the levels of 1155 SomaLogic assayed proteins in CSF and brain tissue based on a GWAS from Yang *et al*.^33^. Significant associations were prioritized based on prior evidence implicating the identified protein for involvement with neurodegeneration. Specifically, GWAS catalog^34^ was queried to identify protein encoding genes associating with traits such as dementia, cognition, brain structure and volume or amyloid plaque accumulation. British National Formulary (BNF, accessed 04-09-2021) and ChEMBL (version 28) were queried to identify compound with affinity for the identified proteins and a known indication or side-effect for neurodegenerative diseases such as dementia, AD or PD^35^. We leveraged Ensembl (GRCh37 Release 110) to identify all gene name synonyms and used these to perform a PubMed query using the following disease relevant keywords: “alzheimer*” [tiab] OR “lewy body” [tiab] OR “dementia*” [tiab]. Proteins with an above averaged research interest were identified using asymptotic Poisson tests evaluating deviation for the average citation number; see Supplementary Table S1. Finally, for the subset of CETP implicated CSF/brain proteins, we sought to determine whether CETP may affect these proteins already in blood plasma. For this we conducted a *cis*-MR analysis using CETP concentration against Somalogic plasma proteins available from Ferkingstad *et al.*^36^

### Effect estimates and multiple testing

Effect estimates are presented in the CETP lowering direction, for the CETP and Apo-B weighted MR analyses this implies the results are presented towards decreasing direction, while the Apo-A1 weighted analyses are presented in the increasing direction. Results are provided with 95% confidence intervals (CI) and p-values. Statistical significance was determined by comparing the p-values against a multiplicity corrected threshold of 0.05/26 ≈ 1.8×10^-3^ for the main analysis focussing on associations with biomarkers and disease, with a p-value threshold of 4.3×10^-5^ employed for the more exploratory analysis of the association between plasma CETP concentration and the levels of 1155 proteins measured in brain and/or CSF. Furthermore, results of the main analysis were replicated by identifying significant and directionally concordant results using the *cis*-MR analysis of CETP activity weighted by Apo-A1 concentration and Apo-B concentration.

## Results

*Cis*-MR was employed to evaluate the potential causal effects lower CETP had on biomarker and disease traits. Specifically, we sourced instruments from a ±25 kbp window within and around *CETP* (ENSG00000087237), selecting variants based on GWAS’ of CETP concentration (no. participants: 4,248), Apo-A1 concentration (no. participants: 355,729), or Apo-B concentration (no. participants: 355,729).

### Effects of lower CETP concentration on biomarker traits

Lower CETP concentration(Figure 1, Table S2) was associated with a decrease in plasma concentration of LDL-C (-0.08 standard deviation (SD), 95%CI -0.09; -0.08), intermediate-density lipoprotein cholesterol (IDL-C: -0.04 SD, 95%CI -0.05; -0.03), very low-density lipoprotein cholesterol (VLDL-C: -0.24 SD, 95%CI -0.27; -0.21), remnant-cholesterol (-0.16 mmol/L, 95%CI -0.17; -0.15, representing the sum of IDL-C and VLDL-C), non-HDL-C (-0.11 SD, 95%CI -0.12; -0.11, representing HDL-C subtracted from total cholesterol), total triglycerides (-0.09 SD, 95%CI -0.09; -0.08), Apo-B (-0.14 SD, 95%CI -0.15; -0.14), and Lp[a] (-1.62 nmol/L, 95%CI -1.91; -1.32). Following the canonical CETPi effects, genetically instrumented lower CETP increased the concentration of HDL-C (0.59 SD, 95%CI 0.57; 0.60), and Apo-A1 (0.48 SD, 95%CI 0.45; 0.52), respectively.

**Figure 1.**
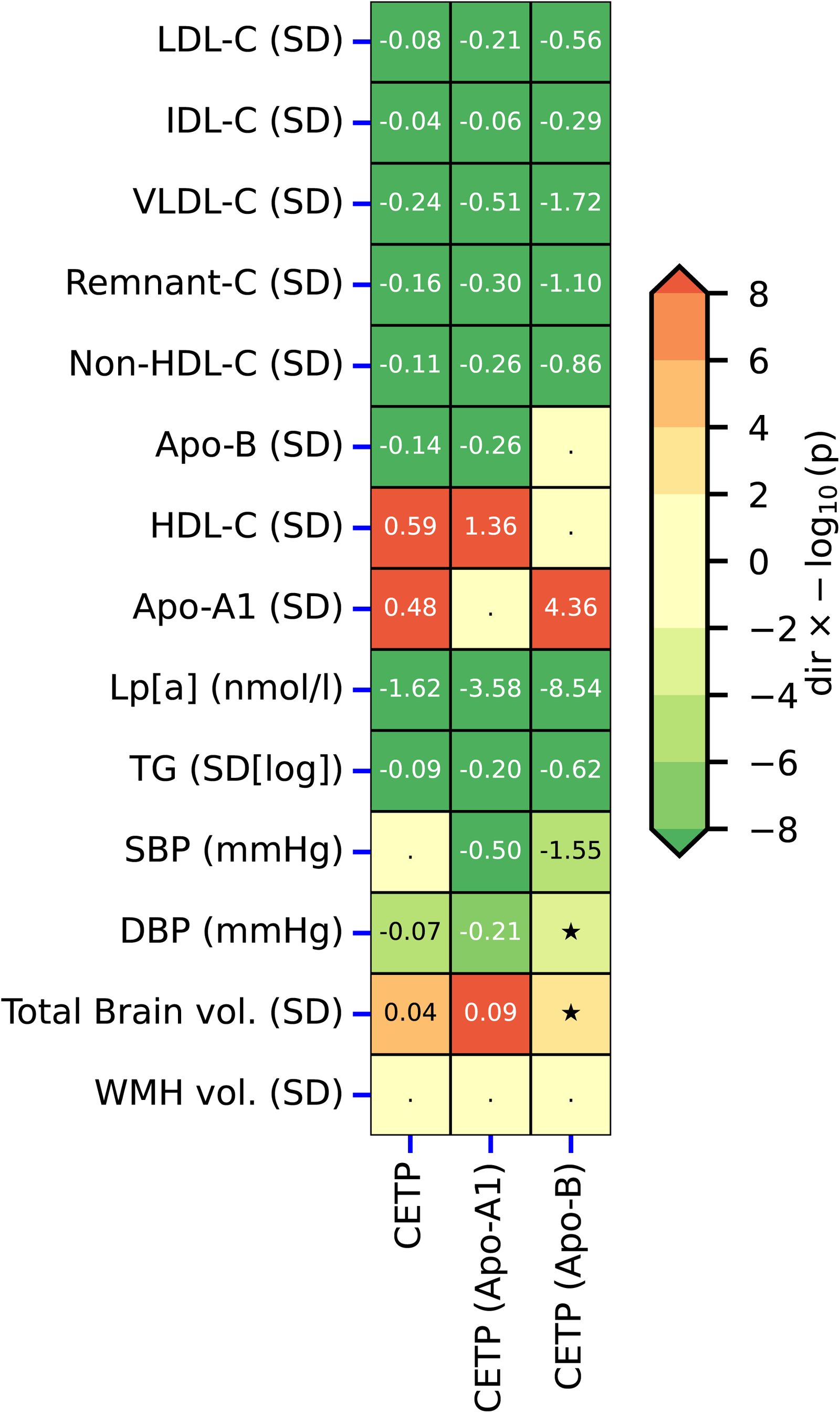
Biomarker effects of lower CETP activity estimated through *cis* Mendelian randomization using three distinct weighting strategies. The MR effects were estimated by alternatingly selecting instruments based on the genetic association with lower CETP concentration (µg/mL), higher Apolipoprotein-A1 (Apo-A1 in g/L), and lower Apolipoprotein-B (Apo-B in g/L). Results are presented as effect direction multiplied by the -log10(p-value), truncated to a maximum of 8. Results with a p-value smaller than 0.05/26 are annotated by the point estimates rounded to two decimal places, nominal significance with a p-value between 0.05 and 0.05/26 is indicated by a star symbol. For the Apo-A1 weighted analyses, the Apo-A1 association was removed (reflecting identical data), with similar masking for the Apo-B weighted analysis. Abbreviations: LDL-C, low-density lipoprotein cholesterol; IDL-C, intermediate-density lipoprotein cholesterol; very low-density lipoprotein cholesterol; HDL-C, high-density lipoprotein cholesterol; Lp[a], lipoprotein a; TG, total triglycerides; SBP, systolic blood pressure; DBP, diastolic blood pressure; vol., volume. Please see Supplementary Table S2 for the full results.

CETP additionally affected non-lipid traits, where lower CETP concentration decreased diastolic blood pressure (DBP) -0.07 mmHg (95%CI -0.11; -0.03), and increased total brain volume 0.04 SD (95%CI 0.02; 0.06), respectively. The presented results were replicated in MR analyses selecting and weighting *CETP* genetic instruments by Apo-A1 and/or Apo-B concentration as a proxy for reduced CETP activity; Figure 1.

### Effects of lower CETP concentration on cardiovascular outcomes

We confirmed that lower plasma CETP concentration decreased the risk of CHD (OR 0.92, 95%CI 0.89; 0.96, p-value 2.47×10), any stroke (OR 0.90, 95%CI 0.85; 0.95), any ischemic stroke (OR 0.96, 95%CI 0.94; 0.98), as well as small vessel stroke (OR 0.90, 95%CI 0.85; 0.96); Figure 2, Table S2. We additionally observed that lower plasma concentration of CETP decreased the risk of AAA (OR 0.76, 95%CI 0.73; 0.80) and HF (OR 0.97, 95%CI 0.93; 1.00, p-value 4.39×10^⁻²^), although the latter only reached nominal significance. Aside from the stroke and ischemic stroke signals, which were partially replicated by Apo-A1 weighted MRs, the associations with CHD, small vessel stroke, HF and AAA were fully replicated by *cis*-MR analyses selecting and weighting *CETP* variants by their associations on Apo-A1 or Apo-B concentration; Figure 2.

**Figure 2.**
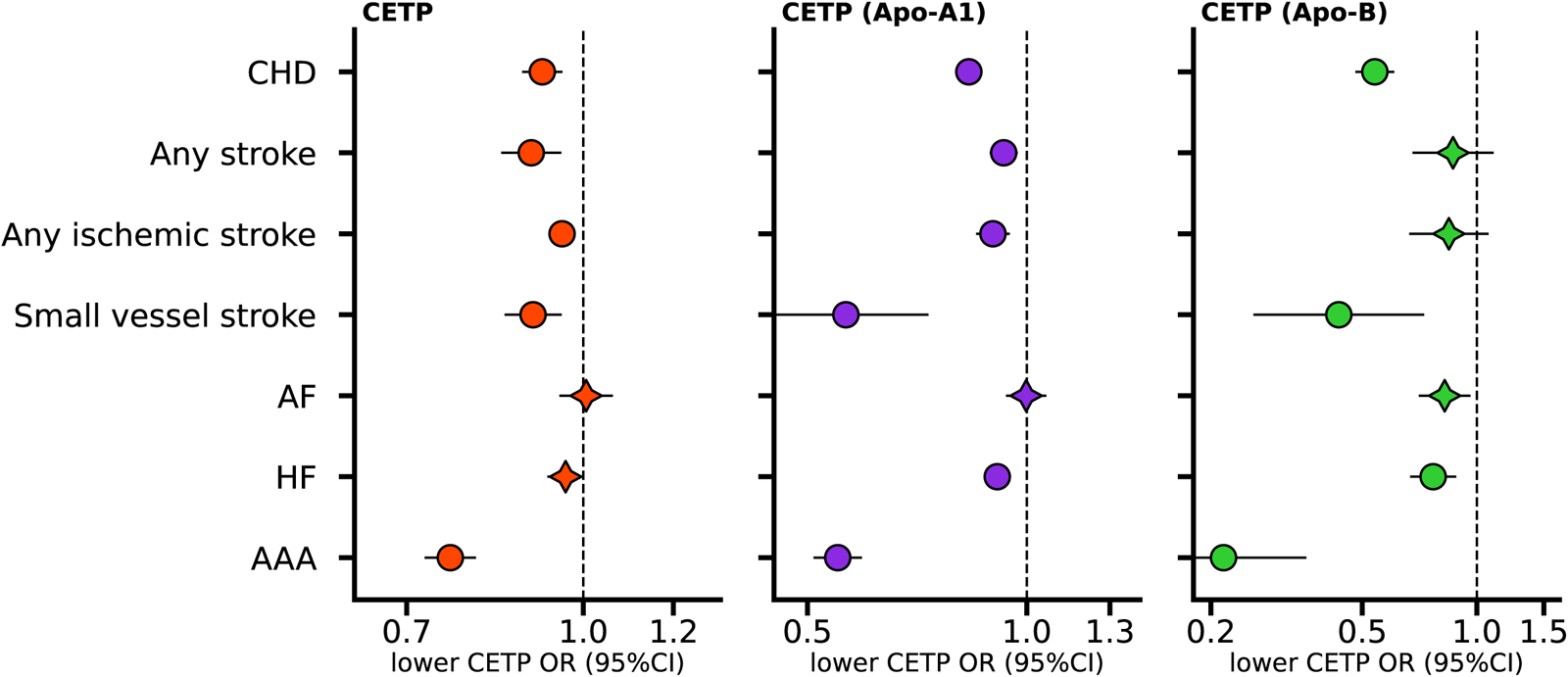
Cardiovascular effects of lower CETP activity on cardiovascular disease estimated through *cis* Mendelian randomization using three distinct weighting strategies. The MR effects are estimated by alternatingly selecting instruments based on the genetic association with lower CETP concentration (µg/mL), higher Apolipoprotein-A1 (Apo-A1 in g/L), and lower Apolipoprotein-B (Apo-B in g/L). The estimated odds ratio (OR) is indicated by a circle if the p-value was smaller than 0.05/26, or by a star otherwise, with the horizontal bars representing 95% confidence intervals (95%CI), a neutral effect of 1 is indicated by the dashed vertical line. Abbreviations: CHD, coronary heart disease; AF, atrial fibrillation; HF, heart failure; AAA, abdominal aortic aneurysm. Please see Supplementary Table S2 for the full results.

### Effects of lower CETP concentration on neurodegenerative outcomes

Noting that lower plasma concentration of CETP associated with higher brain volume, we next explored associations with neurodegenerative traits. We found that lower CETP concentration was associated with a decrease in LBD (OR 0.81, 95%CI 0.74; 0.89, p-value 2.95×10), where *APOE4-* ε4 *status* modified this association: LBD in *APOE-*ε4 carriers (OR 0.61, 95%CI 0.51; 0.73, p-value 4.91×10),and non *APOE-*ε4 carriers (OR 0.89, 95%CI 0.79; 1.01, p-value 8.06×10^⁻²^); interaction p-value 5.81×10^-4^. We further observed that lower CETP concentration protected against dementia in Parkinson’s disease (OR 0.26, 95%CI 0.14; 0.48, p-value 1.29×10), which partially overlaps with known LBD pathophysiology; Figure 3, Table S2. Additionally, we observed a nominal risk decreasing effect of lower CETP on ALS (OR 0.85, 95%CI 0.75; 0.97, p-value 1.64×10^⁻²^). The apolipoprotein (both Apo-A1 and Apo-B) weighted analyses replicated the LBD in *APOE-*ε4 carriers, with the Apo-A1 weighted analysis also replicating the associations for dementia in PD, as well as the ALS association; Figure 3.

**Figure 3.**
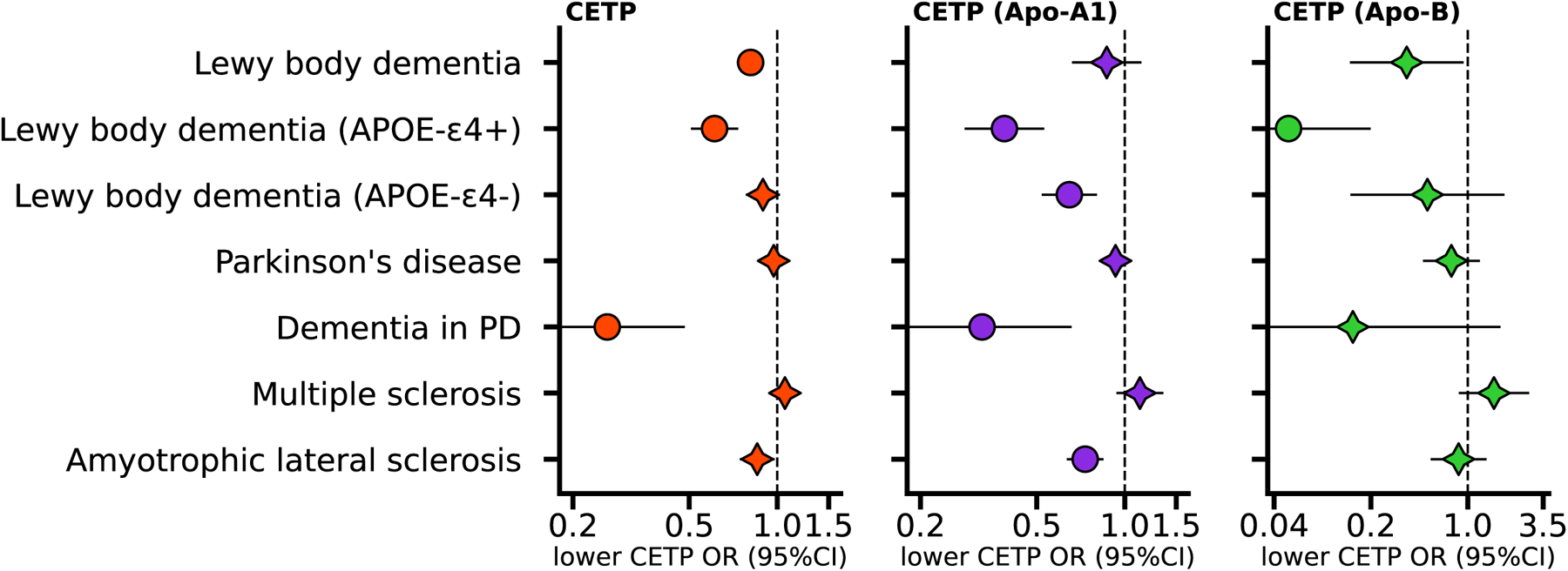
Effects of lower CETP activity on neurological traits estimated through *cis* Mendelian randomization using three distinct weighting strategies. The MR effects are estimated by alternatingly selecting instruments based on the genetic association with lower CETP concentration (µg/mL), higher Apolipoprotein-A1 (Apo-A1 in g/L), and lower Apolipoprotein-B (Apo-B in g/L). The estimated odds ratio (OR) is indicated by a circle if the p-value was smaller than 0.05/26, or by a star otherwise, with the horizontal bars representing 95% confidence intervals (95%CI), a neutral effect of 1 is indicated by the dashed vertical line. Abbreviations: PD, Parkinson’s disease; MS. Note that *APOE-*ε4+ refers to ε4 carriers, *APOE-*ε4-refers to non-carriers. Please see Supplementary Table S2 for the full results.

### Association of plasma CETP concentration with protein values in brain and CSF

We identified (Figure 4, Supplementary Tables S3-S4) associations between lower plasma CETP concentrations and the levels of 26 proteins previously implicated with neurodegenerative traits. This set included the LRP1B encoding gene, whose variants showed an *APOE-* ε4 dependent association with dementia in PD^37^. We additionally found that lower CETP was associated with lower CSF values of EPHA2 and CSF-1, which are targeted by inhibiting compounds in clinical testing for AD. Similarly, we observed that lower CETP increased the values of brain HSP 60^38^ and CSF FAM3B^39^, which are anticipated to prevent β-amyloid aggregation and generation. Where FAM3B was also affected by CETP in plasma (Table S5).

**Figure 4.**
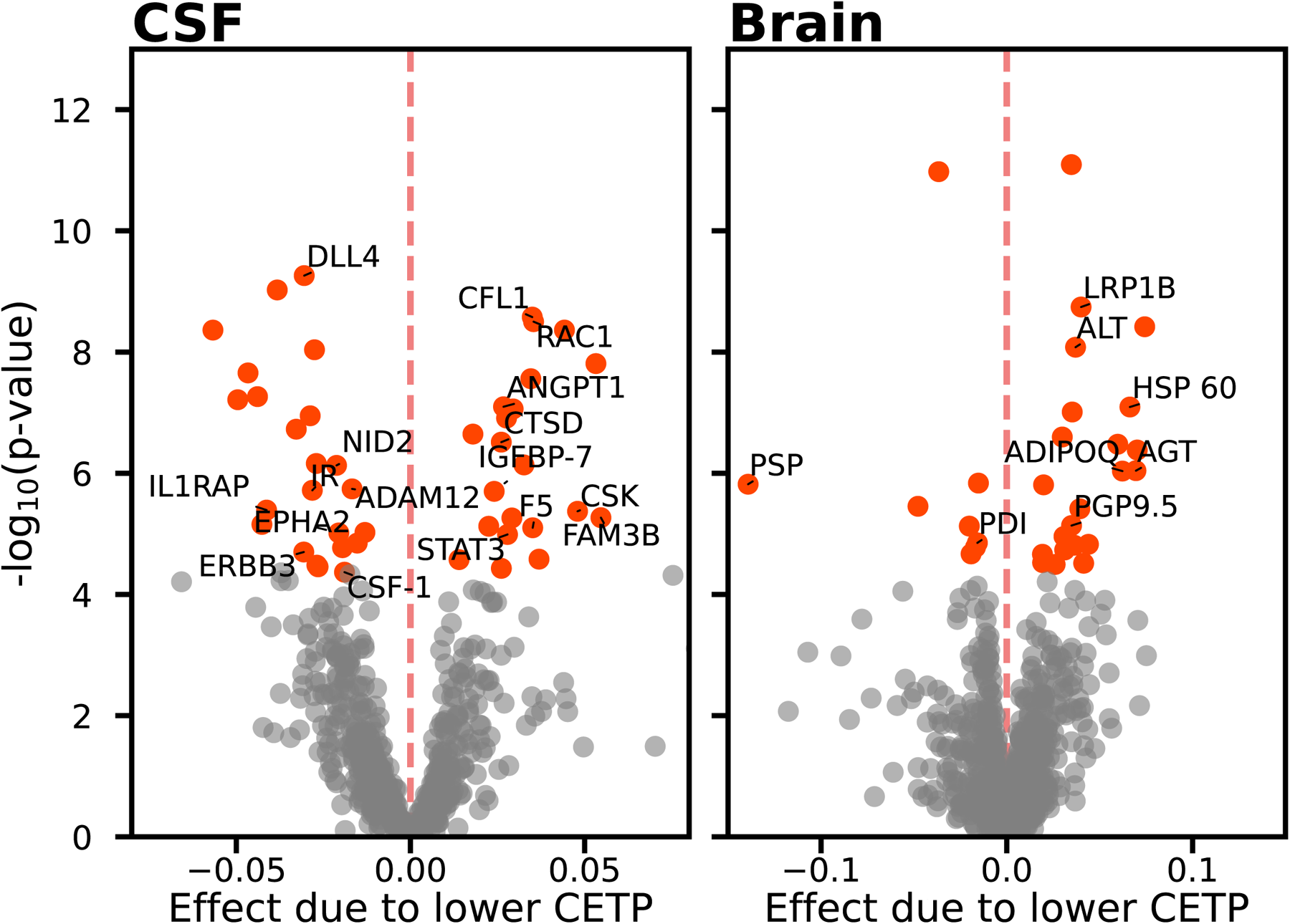
The association of lower plasma CETP with protein values in CSF or brain tissue. Estimates are derived from a *cis* MR analyses of lower plasma concentration exploring associations with SomaLogic derived proteins expressed in Cerebrospinal fluid (CSF) or brain tissue^33^. Estimates are provided for a µg/mL decrease in CETP plasma concentration as mean difference in standard deviation of protein value is plotted on the x-axis, against the -log_10_(p-value) on the y-axis. Associations passing a multiplicity corrected p-value of 4.3×10^-5^ are depicted in orange, with proteins enriched for neurodegeneration annotated by ensembl gene name. Neurodegeneration enrichment was based on information from GWAS catalog, automated PubMed queries, and BNF and ChEMBL providing information on known neurological indications or side-effects for drug compounds targeting these proteins. See Supplementary Tables S1, S3-S4.

## Discussion

In the current analysis we employed *cis*-MR to determine biological consequences of lower CETP activity. We recapitulated and extended known beneficial effects of lower CETP levels on blood lipids, as well as protective effects on cardiovascular diseases such as CHD, AAA, HF, and small vessel stroke. Subsequently we explored potential associations with dementia related traits, noting protective effects of lower CETP concentrations with LBD which was most pronounced in *APOE-*ε4 carriers – compatible with the previously observed protective effect of loss-of-function CETP variants in *APOE-*ε4 carriers^40^. The effects of lower CETP concentration were replicated by performing *cis*-MR weighting *CETP* variants by their association on Apo-A1 and Apo-B concentration. The protective effect on LBD was further supported by a protective effect of lower CETP concentration on dementia in people with PD, reflecting underlying shared aetiology due to Lewy body disorders^17^. We additionally, observed that lower plasma concentrations of CETP were associated with protein pathways in CSF and the brain, some of which are already in clinical phase testing for AD treatment, linked to neurodegeneration through *APOE-*ε4 dependent lipid metabolism and β-amyloid aggregation and degradation.

The observed association between low activity of CETP and dementia is in-line with our current understanding of lipid metabolism, whereby the lower LDL-C levels, higher levels of HDL particles and increased circulating Apo-A1 levels, might protect against the *APOE4-*ε4 dependent effects on cholesterol transport, and on oxysterol and β-amyloid clearance. It is worth emphasizing that the LBD GWAS^41,42^ we used, uniquely performed autopsy-confirmed case-ascertainment, hence the observed associations reflect true LBD rather than a diagnosis based on clinical manifestation. Importantly, we did observe a protective, albeit relatively attenuated, effect of lower CETP concentration decreasing the risk of LBD in non *APOE-*ε4 carriers when weighting by Apo-A1: OR 0.65 95%CI 0.52; 0.80 (p-value 4.11×10). This association did not reach statistical significance in the Apo-B or CETP concentration weighted analyses and therefore requires further confirmation. Given the observed *APOE4* interaction with CETP, and the absence of *APOE4* stratified GWAS we did not explore associations with AD. Furthermore, while we did observed replicated effects of lower CETP concentration on larger brain volume, we did not observe a similarly concordant effect of lower CETP on white matter hyperintensity volume. Potentially, this reflects the lack of *APOE*4 stratification^43^, lack of data on regional volumes^44^, or simply distinct pathways with brain volume more closely relating to plaque forming. Additionally, we found a protective effect of lower CETP against dementia in PD. Furthermore, *APOE-*ε4 carriership increases the risk of both PB, LBD, and dementia in PD^45^, hence this observed association provided further justification for *APOE4* modifying the effects of CETP on neurodegenerative traits.

As described previously^9,11^ a *cis-*MR analysis weighted by a downstream biomarker which is affected by the protein provides inference on the protein effect direction conditional on firm understanding on whether the protein increases or decreases biomarker concentration and/or activity. Such a biomarker weighted MR analysis does not however provide evidence of potential downstream mediation effects. As such the presented Apo-A1 and Apo-B weighted *cis-*MR analysis represents directional tests of the effect of CETP *activity*, not of potential mediation by either apolipoproteins. Given the distinction between protein concentration and activity, the Apo-A1 and Apo-B weighted analyses – representing a combination of CETP concentration and activity rather than concentration alone, not only serve as partial replication, but also complements the *cis*-MR analysis based on CETP concentration. While the current analyses suggest that inhibition of CETP might protect against dementia, it does not provide information on the required dosage, timing and duration of CETP inhibition^8^. As such the reported effect estimates, while robust indicators of effect direction, are unlikely to reflect anticipated effect magnitudes of pharmacological inhibition of CETP. Our findings therefore call for careful re-analysis of existing (pre) clinical data on CETP inhibition, followed by potential *de novo* studies evaluating potential effects of CETP inhibition on dementia. In fact, our analyses are supported by recent studies in mice transgenic for both the human amyloid precursor protein (*APP*) gene, as well as *CETP*, showing accelerated AD progression concomitant with a 22% increase of cholesterol content in brain^24^. Moreover, administration of the CETPi evacetrapib rescued memory deficit in these *AAP/CETP*tg mice^24^. This beneficial change in cognition in evacetrapib treated mice correlated with both decreased LDL-C as well as increased HDL-C levels conferred by the CETPi.

The MR analyses performed in this study are protected against bias due to pre-translational horizontal pleiotropy by combining a model selection framework (providing a data-driven choice between IVW and MR-Egger MR methods) with removal of potential pleiotropic variants based on contributions to the leverage or heterogeneity statistics^31^. Furthermore, our analysis of the effect of lower CETP concentration on blood lipids and CVD outcomes are in-line with findings from CETP inhibitor trials^5,6^, strongly suggesting the presented MR findings are protected against pre-translational pleiotropy. Given that the effects of CETP on dementia are anticipated to follow from its effect on lipid metabolism, this implies that the associations with dementia traits are similarly robust to pre-translational horizontal pleiotropy bias. Furthermore, our analysis was protected against bias due to potential weak-instruments and winner’s curse by selecting genetic variants strongly related with CETP concentration, using a F-statistic threshold of 24 or larger. Second, the GWAS on CETP concentration was sourced from a single study conducted by Blauw *et al*.^26^, which has no sample overlap with the outcome GWAS ensuring that on average, any potential for weak instrument bias acts towards a neutral effect direction^46^.

In conclusion, our *cis*-MR recapitulated the beneficial on-target effects of lower CETP activity on blood lipids and CVD outcomes, mimicking the effect of pharmacologic CETP-inhibition. Consistent with known pathophysiology we expanded these analyses to show that lower CETP activity may elicit an *APOE4* dependent protective effect on Lewy body dementia and dementia associated with Parkinson’s disease. In conjunction with human data of loss-of-function alleles of CETP that protect against dementia in *APOE-*ε4+ carriers and preclinical data in a humanized rodent model of dementia that show rescue of cognition loss by a CETP-inhibition, these results suggest that CETP-inhibition might be repurposed for treatment of dementia in *APOE-*ε4+ carriers.

## Acknowledgments

A preprint version of this manuscript has been deposited at: <URL available upon publication>. This research has been conducted using the UK Biobank Resource under Application Number 12113. The authors are grateful to UK Biobank participants.

## Author contributions

AFS, CF, JJK designed the study. AFS performed the analyses and drafted the manuscript. MHD, MD, JJK, and CF provided critical input on the analysis, as well as the drafted manuscript.

## Code availability

Analyses were conducted using Python v3.7.13 (for GNU Linux), Pandas v1.3.5, Numpy v1.21.6, bio-misc v0.1.4, and matplotlib v3.4.3.

## Data availability

The genetic data used for this analyses has been deposited on figshare: [URL available upon acceptance]. The individual GWAS data leveraged in this study can be accessed as followed: CETP concentration was available from Blauw *et al.* (n: 4,248 https://www.ahajournals.org/doi/full/10.1161/CIRCGEN.117.002034), Apo-A1, Apo-B, IDL-C, VLDL-C, remnant-C, from (n: 115,078, https://gwas.mrcieu.ac.uk/datasets, dataset: met-d), LDL-C, HDL-C, and TG from (n: 1,320,016, http://csg.sph.umich.edu/willer/public/glgc-lipids2021), lp[a] from (n: 361,194, http://www.nealelab.is/uk-biobank), systolic/diastolic blood pressure (n: 757,601, https://www.ebi.ac.uk/gwas/publications/30224653), brain volume, (n: 47,316, https://ctg.cncr.nl/software/summary_statistics), white matter hyperintensity volume (n: 42,310, https://www.ebi.ac.uk/gwas/publications/32358547), coronary heart disease (cases: 181522, total n:1165690, https://www.ebi.ac.uk/gwas/publications/36474045), any stroke, ischemic stroke, small vessel stroke (any stroke cases: 40,585, total n: 446,696; any ischemic stroke cases: 34,217, total n: 440,328; small vessel stroke cases: 5,386, total n: 411,497, http://www.megastroke.org/index.html), atrial fibrillation (cases: 60,620, total n: 1,030,836, https://www.ebi.ac.uk/gwas/publications/30061737), heart failure (cases: 115,150, total n: 1,665,481, https://www.ebi.ac.uk/gwas/publications/36376295), aortic atrial aneurysm (cases: 8,163, total n:1,164,713, https://www.globalbiobankmeta.org/), Lewy body dementia (cases: 2,981, total n: 6,618, https://www.ebi.ac.uk/gwas/publications/33589841), Lewy body stratified on *APOE-*ε4 status (positive cases: 1,180, positive total n: 1,837, negative cases: 1,286, negative total cases: 3,557, https://www.ebi.ac.uk/gwas/publications/35381062), Parkinson’s disease (cases: 56,306, total n: 14,056,306, https://www.thelancet.com/pdfs/journals/laneur/PIIS1474-4422(19)30320-5.pdf), dementia in Parkinson’s disease (cases: 263, total n: 3,923, https://pdgenetics.org/resources), multiple sclerosis (cases: 14,498, total n:38,589, https://imsgc.net/), amyotrophic lateral sclerosis (cases: 15,156, total n: 41,398, https://www.nature.com/articles/ng.3622).

## References

1. Kastelein JJ, Kereiakes DJ, Cannon CP, Bays HE, Minini P, Lee LV, Maroni J, Farnier M. Additional LDL-C reduction achieved with alirocumab dose increase on background statin. Circulation 2015;132.

2. Nicholls SJ, Ditmarsch M, Kastelein JJ, Rigby SP, Kling D, Curcio DL, Alp NJ, Davidson MH. Lipid lowering effects of the CETP inhibitor obicetrapib in combination with high-intensity statins: a randomized phase 2 trial. Nat Med 2022;28:1672–1678.

3. Ballantyne CM, Ditmarsch M, Kastelein JJ, Nelson AJ, Kling D, Hsieh A, Curcio DL, Maki KC, Davidson MH, Nicholls SJ. Obicetrapib plus ezetimibe as an adjunct to high-intensity statin therapy: A randomized phase 2 trial. J Clin Lipidol 2023:S1933–2874(23)00186-1.

4. Hovingh GK, Kastelein JJP, Deventer SJH van, Round P, Ford J, Saleheen D, Rader DJ, Brewer HB, Barter PJ. Cholesterol ester transfer protein inhibition by TA-8995 in patients with mild dyslipidaemia (TULIP): a randomised, double-blind, placebo-controlled phase 2 trial. The Lancet 2015;386:452–460.

5. Schmidt AF, Hunt NB, Gordillo-Marañón M, Charoen P, Drenos F, Kivimaki M, Lawlor DA, Giambartolomei C, Papacosta O, Chaturvedi N, Bis JC, O’Donnell CJ, Wannamethee G, Wong A, Price JF, Hughes AD, Gaunt TR, Franceschini N, Mook-Kanamori DO, Zwierzyna M, Sofat R, Hingorani AD, Finan C. Cholesteryl ester transfer protein (CETP) as a drug target for cardiovascular disease. Nat Commun 2021;12:5640.

6. The HPS3/TIMI55–REVEAL Collaborative Group. Effects of Anacetrapib in Patients with Atherosclerotic Vascular Disease. N Engl J Med 2017;377:1217–1227.

7. Nicholls SJ, Ray KK, Nelson AJ, Kastelein JJP. Can we revive CETP-inhibitors for the prevention of cardiovascular disease? Curr Opin Lipidol 2022;33:319–325.

8. Cupido AJ, Reeskamp LF, Hingorani AD, Finan C, Asselbergs FW, Hovingh GK, Schmidt AF. Joint Genetic Inhibition of PCSK9 and CETP and the Association With Coronary Artery Disease: A Factorial Mendelian Randomization Study. JAMA Cardiol 2022;7:955–964.

9. Schmidt AF, Finan C, Gordillo-Marañón M, Asselbergs FW, Freitag DF, Patel RS, Tyl B, Chopade S, Faraway R, Zwierzyna M, Hingorani AD. Genetic drug target validation using Mendelian randomisation. Nat Commun 2020;11:3255.

10. Hingorani A, Humphries S. Nature’s randomised trials. Lancet 2005;366:1906–1908.

11. Schmidt AF, Hingorani AD, Finan C. Human Genomics and Drug Development. Cold Spring Harb Perspect Med 2021:a039230.

12. Marsillach J, Adorni MP, Zimetti F, Papotti B, Zuliani G, Cervellati C. HDL Proteome and Alzheimer’s Disease: Evidence of a Link. Antioxidants (Basel) 2020;9:1224.

13. Suidan GL, Ramaswamy G. Targeting Apolipoprotein E for Alzheimer’s Disease: An Industry Perspective. Int J Mol Sci 2019;20:2161.

14. Borràs C, Mercer A, Sirisi S, Alcolea D, Escolà-Gil JC, Blanco-Vaca F, Tondo M. HDL-like-Mediated Cell Cholesterol Trafficking in the Central Nervous System and Alzheimer’s Disease Pathogenesis. Int J Mol Sci 2022;23:9356.

15. Yin F. Lipid metabolism and Alzheimer’s disease: clinical evidence, mechanistic link and therapeutic promise. FEBS J 2023;290:1420–1453.

16. Schmidt AF, Joshi R, Gordillo-Marañón M, Drenos F, Charoen P, Giambartolomei C, Bis JC, Gaunt TR, Hughes AD, Lawlor DA, Wong A, Price JF, Chaturvedi N, Wannamethee G, Franceschini N, Kivimaki M, Hingorani AD, Finan C. Biomedical consequences of elevated cholesterol-containing lipoproteins and apolipoproteins

17. Lippa CF, Duda JE, Grossman M, Hurtig HI, Aarsland D, Boeve BF, Brooks DJ, Dickson DW, Dubois B, Emre M, Fahn S, Farmer JM, Galasko D, Galvin JE, Goetz CG, Growdon JH, Gwinn-Hardy KA, Hardy J, Heutink P, Iwatsubo T, Kosaka K, Lee VM-Y, Leverenz JB, Masliah E, McKeith IG, Nussbaum RL, Olanow CW, Ravina BM, Singleton AB, Tanner CM, Trojanowski JQ, Wszolek ZK, DLB/PDD Working Group. DLB and PDD boundary issues: diagnosis, treatment, molecular pathology, and biomarkers. Neurology 2007;68:812–819.

18. Raulin A-C, Doss SV, Trottier ZA, Ikezu TC, Bu G, Liu C-C. ApoE in Alzheimer’s disease: pathophysiology and therapeutic strategies. Mol Neurodegener 2022;17:72.

19. Strittmatter WJ, Saunders AM, Schmechel D, Pericak-Vance M, Enghild J, Salvesen GS, Roses AD. Apolipoprotein E: high-avidity binding to beta-amyloid and increased frequency of type 4 allele in late-onset familial Alzheimer disease. Proc Natl Acad Sci U S A 1993;90:1977–1981.

20. Lanfranco MF, Ng CA, Rebeck GW. ApoE Lipidation as a Therapeutic Target in Alzheimer’s Disease. Int J Mol Sci 2020;21:6336.

21. Fernández-Calle R, Konings SC, Frontiñán-Rubio J, García-Revilla J, Camprubí-Ferrer L, Svensson M, Martinson I, Boza-Serrano A, Venero JL, Nielsen HM, Gouras GK, Deierborg T. APOE in the bullseye of neurodegenerative diseases: impact of the APOE genotype in Alzheimer’s disease pathology and brain diseases. Mol Neurodegener 2022;17:62.

22. Bedlack RS, Strittmatter WJ, Morgenlander JC. Apolipoprotein E and Neuromuscular Disease: A Critical Review of the Literature. Archives of Neurology 2000;57:1561–1565.

23. Berghoff SA, Gerndt N, Winchenbach J, Stumpf SK, Hosang L, Odoardi F, Ruhwedel T, Böhler C, Barrette B, Stassart R, Liebetanz D, Dibaj P, Möbius W, Edgar JM, Saher G. Dietary cholesterol promotes repair of demyelinated lesions in the adult brain. Nat Commun 2017;8:14241.

24. Oestereich F, Yousefpour N, Yang E, Phénix J, Nezhad ZS, Nitu A, Vázquez Cobá A, Ribeiro-da-Silva A, Chaurand P, Munter LM. The cholesteryl ester transfer protein (CETP) raises cholesterol levels in the brain. J Lipid Res 2022;63:100260.

25. Armstrong NS-, Tanigawa Y, Amar D, Mars N, Benner C, Aguirre M, Venkataraman GR, Wainberg M, Ollila HM, Kiiskinen T, Havulinna AS, Pirruccello JP, Qian J, Shcherbina A, consortium F, Rodriguez F, Assimes TL, Agarwala V, Tibshirani R, Hastie T, Ripatti S, Pritchard JK, Daly MJ, Rivas MA. Genetics of 35 blood and urine biomarkers in the UK Biobank. Nat Genet 2021;53:185–194.

26. Blauw LL, Li-Gao R, Noordam R, Mutsert R de, Trompet S, Berbée JFP, Wang Y, Klinken JB van, Christen T, Heemst D van, Mook-Kanamori DO, Rosendaal FR, Jukema JW, Rensen PCN, Willems van Dijk K. CETP (Cholesteryl Ester Transfer Protein) Concentration: A Genome-Wide Association Study Followed by Mendelian Randomization on Coronary Artery Disease. Circ Genom Precis Med 2018;11:e002034.

27. Burgess S, Thompson SG. Avoiding bias from weak instruments in mendelian randomization studies. Int J Epidemiol 2011;40:755–764.

28. Bowden J, Smith GD, Burgess S. Mendelian randomization with invalid instruments: Effect estimation and bias detection through Egger regression. Int J Epidemiol 2015;44:512–525.

29. Burgess S, Zuber V, Valdes-Marquez E, Sun BB, Hopewell JC. Mendelian randomization with fine-mapped genetic data: Choosing from large numbers of correlated instrumental variables. Genetic Epidemiology 2017;41:714–725.

30. Bowden J, Del Greco M F, Minelli C, Davey Smith G, Sheehan N, Thompson J, Del Greco M F, Minelli C, Davey Smith G, Sheehan N, Thompson J. A framework for the investigation of pleiotropy in two-sample summary data Mendelian randomization. Statistics in medicine 2017;36:1783–1802.

31. Bowden J, Spiller W, Del Greco M F, Sheehan N, Thompson J, Minelli C, Davey Smith G. Improving the visualization, interpretation and analysis of two-sample summary data Mendelian randomization via the Radial plot and Radial regression. Int J Epidemiol 2018;47:1264–1278.

32. Rücker G, Schwarzer G, Carpenter JR, Binder H, Schumacher M. Treatment-effect estimates adjusted for small-study effects via a limit meta-analysis. Biostatistics 2011;12:122–142.

33. Yang C, Farias FHG, Ibanez L, Suhy A, Sadler B, Fernandez MV, Wang F, Bradley JL, Eiffert B, Bahena JA, Budde JP, Li Z, Dube U, Sung YJ, Mihindukulasuriya KA, Morris JC, Fagan AM, Perrin RJ, Benitez BA, Rhinn H, Harari O, Cruchaga C. Genomic atlas of the proteome from brain, CSF and plasma prioritizes proteins implicated in neurological disorders. Nat Neurosci 2021;24:1302–1312.

34. Buniello A, MacArthur JAL, Cerezo M, Harris LW, Hayhurst J, Malangone C, McMahon A, Morales J, Mountjoy E, Sollis E, Suveges D, Vrousgou O, Whetzel PL, Amode R, Guillen JA, Riat HS, Trevanion SJ, Hall P, Junkins H, Flicek P, Burdett T, Hindorff LA, Cunningham F, Parkinson H. The NHGRI-EBI GWAS Catalog of published genome-wide association studies, targeted arrays and summary statistics 2019. Nucleic Acids Res 2019;47:D1005–D1012.

35. Finan C, Gaulton A, Kruger FA, Lumbers RT, Shah T, Engmann J, Galver L, Kelley R, Karlsson A, Santos R, Overington JP, Hingorani AD, Casas JP, Lumbers T, Shah T, Engmann J, Galver L, Kelly R, Karlsson A, Santos R, Overington JP, Hingorani AD, Casas JP. The druggable genome and support for target identification and validation in drug development. Science Translational Medicine 2017;9.

36. Ferkingstad E, Sulem P, Atlason BA, Sveinbjornsson G, Magnusson MI, Styrmisdottir EL, Gunnarsdottir K, Helgason A, Oddsson A, Halldorsson BV, Jensson BO, Zink F, Halldorsson GH, Masson G, Arnadottir GA, Katrinardottir H, Juliusson K, Magnusson MK, Magnusson OT, Fridriksdottir R, Saevarsdottir S, Gudjonsson SA, Stacey SN, Rognvaldsson S, Eiriksdottir T, Olafsdottir TA, Steinthorsdottir V, Tragante V, Ulfarsson MO, Stefansson H, Jonsdottir I, Holm H, Rafnar T, Melsted P, Saemundsdottir J, Norddahl GL, Lund SH, Gudbjartsson DF, Thorsteinsdottir U, Stefansson K. Large-scale integration of the plasma proteome with genetics and disease. Nat Genet 2021;53:1712–1721.

37. Real R, Martinez-Carrasco A, Reynolds RH, Lawton MA, Tan MMX, Shoai M, Corvol J-C, Ryten M, Bresner C, Hubbard L, Brice A, Lesage S, Faouzi J, Elbaz A, Artaud F, Williams N, Hu MTM, Ben-Shlomo Y, Grosset DG, Hardy J, Morris HR. Association between the LRP1B and APOE loci and the development of Parkinson’s disease dementia. Brain 2023;146:1873–1887.

38. Tang Z, Su K-H, Xu M, Dai C. HSF1 physically neutralizes amyloid oligomers to empower overgrowth and bestow neuroprotection. Sci Adv 2020;6:eabc6871.

39. Hasegawa H, Liu L, Tooyama I, Murayama S, Nishimura M. The FAM3 superfamily member ILEI ameliorates Alzheimer’s disease-like pathology by destabilizing the penultimate amyloid-β precursor. Nat Commun 2014;5:3917.

40. Chemparathy A, Guen YL, Chen S, Lee E-G, Leong L, Gorzynski J, Xu G, Belloy M, Kasireddy N, Tauber AP, Williams K, Stewart I, Wingo T, Lah J, Jayadev S, Hales C, Peskind E, Child DD, Keene CD, Cong L, Ashley E, Yu C-E, Greicius MD. APOE loss-of-function variants: Compatible with longevity and associated with resistance to Alzheimer’s Disease pathology. medRxiv 2023:2023.07.20.23292771.

41. Chia R, Sabir MS, Bandres-Ciga S, Saez-Atienzar S, Reynolds RH, Gustavsson E, Walton RL, Ahmed S, Viollet C, Ding J, Makarious MB, Diez-Fairen M, Portley MK, Shah Z, Abramzon Y, Hernandez DG, Blauwendraat C, Stone DJ, Eicher J, Parkkinen L, Ansorge O, Clark L, Honig LS, Marder K, Lemstra A, St George-Hyslop P, Londos E, Morgan K, Lashley T, Warner TT, Jaunmuktane Z, Galasko D, Santana I, Tienari PJ, Myllykangas L, Oinas M, Cairns NJ, Morris JC, Halliday GM, Van Deerlin VM, Trojanowski JQ, Grassano M, Calvo A, Mora G, Canosa A, Floris G, Bohannan RC, Brett F, Gan-Or Z, Geiger JT, Moore A, May P, Krüger R, Goldstein DS, Lopez G, Tayebi N, Sidransky E, American Genome Center, Norcliffe-Kaufmann L, Palma J-A, Kaufmann H, Shakkottai VG, Perkins M, Newell KL, Gasser T, Schulte C, Landi F, Salvi E, Cusi D, Masliah E, Kim RC, Caraway CA, Monuki ES, Brunetti M, Dawson TM, Rosenthal LS, Albert MS, Pletnikova O, Troncoso JC, Flanagan ME, Mao Q, Bigio EH, Rodríguez-Rodríguez E, Infante J, Lage C, González-Aramburu I, Sanchez-Juan P, Ghetti B, Keith J, Black SE, Masellis M, Rogaeva E, Duyckaerts C, Brice A, Lesage S, Xiromerisiou G, Barrett MJ, Tilley BS, Gentleman S, Logroscino G, Serrano GE, Beach TG, McKeith IG, Thomas AJ, Attems J, Morris CM, Palmer L, Love S, Troakes C, Al-Sarraj S, Hodges AK, Aarsland D, Klein G, Kaiser SM, Woltjer R, Pastor P, Bekris LM, Leverenz JB, Besser LM, Kuzma A, Renton AE, Goate A, Bennett DA, Scherzer CR, Morris HR, Ferrari R, Albani D, Pickering-Brown S, Faber K, Kukull WA, Morenas-Rodriguez E, Lleó A, Fortea J, Alcolea D, Clarimon J, Nalls MA, Ferrucci L, Resnick SM, Tanaka T, Foroud TM, Graff-Radford NR, Wszolek ZK, Ferman T, Boeve BF, Hardy JA, Topol EJ, Torkamani A, Singleton AB, Ryten M, Dickson DW, Chiò A, Ross OA, Gibbs JR, Dalgard CL, Traynor BJ, Scholz SW. Genome sequencing analysis identifies new loci associated with Lewy body dementia and provides insights into its genetic architecture. Nat Genet 2021;53:294–303.

42. Kaivola K, Shah Z, Chia R, Scholz SW. Genetic evaluation of dementia with Lewy bodies implicates distinct disease subgroups. Brain 2021;145:1757–1762.

43. Mirza SS, Saeed U, Knight J, Ramirez J, Stuss DT, Keith J, Nestor SM, Yu D, Swardfager W, Rogaeva E, St. George Hyslop P, Black SE, Masellis M. APOE ε4, white matter hyperintensities, and cognition in Alzheimer and Lewy body dementia. Neurology 2019;93:e1807–e1819.

44. Lorenzini L, Ansems LT, Lopes Alves I, Ingala S, Vállez García D, Tomassen J, Sudre C, Salvadó G, Shekari M, Operto G, Brugulat-Serrat A, Sánchez-Benavides G, Kate M ten, Tijms B, Wink AM, Mutsaerts HJMM, Braber A den, Visser PJ, Berckel BNM van, Gispert JD, Barkhof F, Collij LE. Regional associations of white matter hyperintensities and early cortical amyloid pathology. Brain Commun 2022;4:fcac150.

45. Pankratz N, Byder L, Halter C, Rudolph A, Shults CW, Conneally PM, Foroud T, Nichols WC. Presence of an APOE4 allele results in significantly earlier onset of Parkinson’s disease and a higher risk with dementia. Mov Disord 2006;21:45–49.

46. Burgess S, Davies NM, Thompson SG. Bias due to participant overlap in two-sample Mendelian randomization. 2016;40:597–608.

